# The wide spectrum of neuropsychiatric complications in Covid-19 patients within a multidisciplinary hospital context

**DOI:** 10.1101/2020.10.21.20216747

**Authors:** Cécile Delorme, Marion Houot, Charlotte Rosso, Stéphanie Carvalho, Thomas Nedelec, Redwan Maatoug, Victor Pitron, Salimata Gassama, Sara Sambin, Stéphanie Bombois, Bastien Herlin, Gaëlle Ouvrard, Gaëlle Bruneteau, Adèle Hesters, Ana Zenovia Gales, Bruno Millet, Foudil Lamari, Stéphane Lehericy, Vincent Navarro, Benjamin Rohaut, Sophie Demeret, Thierry Maisonobe, Marion Yger, Bertrand Degos, Louise-Laure Mariani, Christophe Bouche, Nathalie Dzierzynski, Bruno Oquendo, Flora Ketz, An-Hung Nguyen, Aurélie Kas, Jean-Yves Delattre, Jean-Christophe Corvol, on behalf of the CoCo-Neurosciences Study Group

## Abstract

**Objective:** To describe the spectrum of neurological and psychiatric complications in patients with Covid-19 seen in a multidisciplinary center over six months.

**Methods:** We conducted a retrospective, observational study on all patients showing neurological or psychiatric symptoms in the context of Covid-19 seen in the Department of Neurology and Psychiatry of the APHP-Sorbonne University. We collected demographic data, medical and treatment history, comorbidities, symptoms, date of onset, and severity of Covid-19 infection, neurological and psychiatric symptoms, neurological and psychiatric examination data and, when available, results from cerebrospinal fluid (CSF) analysis, brain magnetic resonance (MRI) imaging, 18-fluorodesoxyglucose-position emission computed tomography (FDG-PET/CT)), electroencephalography (EEG) and electroneuromyography (ENMG).

**Results:** 245 patients were included in the analysis. One-hundred fourteen patients (47%) were admitted to the intensive care unit (ICU) and 10 (4%) died. The most frequently reported neuropsychiatric symptoms were motor deficit (41%), cognitive disturbance (35%), impaired consciousness (26%), psychiatric disturbance (24%), headache (20%) and behavioral disturbance (18%). The most frequent syndromes diagnosed were encephalopathy (43%), critical illness polyneuropathy and myopathy (26%), isolated psychiatric disturbance (18%), and cerebrovascular disorders (16%). No patients showed evidence of SARS-CoV-2 in their CSF. Encephalopathy was associated with greater age and higher risk of death. Critical illness neuromyopathy was associated with an extended stay in the ICU.

**Conclusions:** The majority of the neuropsychiatric complications recorded could be imputed to critical illness, intensive care and systemic inflammation, which contrasts with the paucity of more direct SARS-CoV-2-related complications or post-infection disorders.

## Introduction

Covid-19, caused by the SARS-CoV-2 virus, has spread since December 2019 and was soon declared a pandemic by the World Health Organization (WHO). An initial Chinese cohort of 214 Covid-19 patients reported a high frequency of neurological symptoms (36%), including non-specific manifestations such as headache, confusion and myalgia, but also strokes and seizures. A French study in 58 intensive-care unit (ICU) patients further reported a high frequency of encephalopathy and corticospinal signs ^1^. Since then, a variety of neurological complications have been described, including cerebrovascular complications ^2–5^, encephalitis^6,7^, acute disseminated encephalomyelitis (ADEM) ^8^, seizures ^9,10^, myelitis ^11^, Guillain-Barré syndrome ^12^, cranial nerve palsies ^13,14^, anosmia and dysgeusia ^15–17^. Several studies have raised concerns about a high prevalence of anxiety, mood disorders and post-traumatic stress disorders in Covid-19 patients ^18^.

Most of our knowledge of neuropsychiatric complications of Covid-19 derives from case reports and case series. Studies presenting a wider picture of these complications and of their relative frequency are lacking ^19^.

In France, the Paris area has been particularly hit by the outbreak with more than 18,000 patients hospitalized and 6,000 deaths in March and April 2020. The Department of Neurology and Psychiatry of the *Assistance Publique Hôpitaux de Paris (APHP) – Sorbonne University* has been involved in the care of patients affected by Covid-19 and showing neurological or psychiatric symptoms.

Here, we describe the detailed spectrum of neuropsychiatric disorders in 245 Covid-19 patients seen in the Department of Neurology and Psychiatry of APHP - Sorbonne University Hospitals over a six-month period.

## Methods

### Study design and population

This is a retrospective, observational study conducted consecutively on all patients with neurological or psychiatric symptoms in the context of Covid-19 seen between March 1^st^ and August 28^th^ at the *APHP– Sorbonne University*, which groups five University teaching Hospitals in the East of Paris (Pitié-Salpêtrière, Saint-Antoine, Tenon, Rothschild, Charles-Foix hospitals). Investigators were physicians from the Department of Neurology and Psychiatry, which includes all medical units in the field of adult neurology, neurovascular, neurological intensive care, neurorehabilitation, psychiatry, neurophysiology, and neuropathology of these hospitals. During the outbreak, neurologists and psychiatrists were directly in charge of patients with Covid-19 showing neuropsychiatric complications, either hospitalized (in their own units or other units such as pneumology, infectious diseases, and ICU) or as outpatients. Geriatricians, rehabilitation doctors and neuroradiologists were also involved in reporting cases.

All consecutive in- or out-patients, aged 18 years or older, with Covid-19 and *de novo* neurological or psychiatric symptoms were included in the study. Patients showing uniquely anosmia and/or dysgeusia were not included. Exclusion criteria were patients who refused the use of their data, and prisoners.

The primary objective of the study was to describe the full spectrum of neurological and psychiatric complications in Covid-19 patients.

### Data collection

Data was collected retrospectively by investigators from medical records and entered into a structured case report form. Items collected included demographic data, medical and treatment history, comorbidities, symptoms, date of onset, and severity of Covid-19 infection, neurological and psychiatric symptoms, neurological and psychiatric examination, and, when available, results from cerebrospinal fluid (CSF) analysis, brain imaging (computed tomography (CT), magnetic resonance imaging (MRI), 18-fluorodesoxyglucose-positron emission computed tomography (FDG-PET/CT)), electroencephalography (EEG) and electroneuromyography (ENMG).

Covid-19 was defined by at least one of the three following criteria: a) positive SARS-CoV-2 polymerase chain reaction (PCR) in swab or upper pulmonary samples, or positive serology; b) thoracic radiological findings typical of SARS-CoV2 infection in the context of pandemic outbreak; c) suspected Covid-19 infection according to WHO guidance criteria ^20^. The severity of Covid-19 was the status of the patient at the nadir of the disease according to the seven levels as defined by the WHO: 1-not hospitalized, no limitation in daily life activity; 2-not hospitalized, with limitation in daily life activity; 3-hospitalized, no oxygen requirement; 4-hospitalized, necessitating oxygen; 5-hospitalized, necessitating non-invasive ventilation or Optiflow™; 6-hospitalized, necessitating intubation or extracorporeal membrane oxygenation (ECMO); 7-death.

The investigators completed the database consisting of a predefined list of neurological and psychiatric characteristics (symptoms, clinical signs, date of onset). Each item was scored as present, absent or unknown (no clinical evaluation available), and a final neurological or psychiatric syndrome was determined. One patient may have had one or more neurological or psychiatric diagnosis during Covid-19. One investigator (C.D.) reviewed all cases, and classified the diagnoses according to criteria of the Liverpool Brain infections Group (Neuro Network) ^21^. When a patient had more than one diagnosis, the diagnoses were classified as primary (pronounced) or secondary. In the case of conflicting diagnoses, the investigator who entered the case data was contacted to reach consensus.

### Standard Protocol Approvals

The study was conducted in accordance with good clinical practice, the French regulation for retrospective studies on clinical data, and was compliant with the European General Data Protection Regulation (GDPR) and the French *Commission Nationale de l’Informatique et des Libertés* (CNIL) rules. All patients (or their relatives in cases of impaired consciousness) received written information about the research, and consented to the use of their data. The study received the approval of the Sorbonne University Ethics Committee (N°2020 CER-202028). The study is registered on the clinicaltrials.gov website (NCT04362930).

### Statistical analysis

For the analyses, we grouped the 7-level Covid-19 severity score into four categories: 1-not hospitalized (levels 1 and 2); 2-hospitalized without intensive care (levels 3 and 4); 3-hospitalized with intensive care (levels 5 and 6); 4-death (level 7). Kruskal-Wallis tests and Fisher’s Exact tests were used to compare overall differences between groups in demographics and clinical data. Pairwise comparisons were performed using pairwise Wilcoxon–Mann–Whitney tests and pairwise Fisher’s Exact tests with Benjamini-Hochberg method to correct for multiple testing.

In order to investigate the effects of simultaneous risk factors of Covid-19 severity, we performed adjacent category ordinal logistic regression on the four grouped categories of the 7-levels score. Risk factors were age, gender, presence of at least one comorbidity, and presence of obesity.

Presence of neuropsychiatric symptoms, clinical examination features and syndromes were assessed on non-missing data.

For the correlation matrix, we calculated the Pearson correlation for inter-relationships between the following variables: age, gender, presence of one comorbidity, ICU hospitalization, and the various neurological syndromes. Hierarchical clustering was applied with the single linkage distance.

The risk factors for the most frequent neurological syndromes were analyzed using the Classification And Regression Tree (CART) algorithm. The CART algorithm, also known as a “decision tree”, is a non-parametric supervised technique that combines variables in such a way as to best discriminate two groups. For each neurological syndrome, we trained one tree to depth 4 through entropy minimization. The trees are purely descriptive. Statistical analyses were performed using R 3.6.1. and package VGAM_(version 1.1-3) for the ordinal logistic regression (R Foundation for Statistical Computing, Vienna, Austria. URL https://www.R-project.org/.) and using python 3.8 with the scikit learn 0.23.2 package for decision trees and correlation matrices ^22^.

### Data Availability

Anonymized data is available upon request to the corresponding author by request from any qualified investigator.

## Results

### Patient characteristics

During the study period, 1,979 patients were admitted with a diagnosis of confirmed Covid-19 in our center. A total of 249 (12%) Covid-19 patients with a *de novo* neurological or psychiatric manifestation were included in the database. Three patients were excluded because they withdrew consent for the use of their data, and one patient was excluded because he did not fulfill the diagnosis criteria for Covid-19 after careful case review. The population analyzed consisted of 245 cases.

The characteristics of the patients are presented in Table 1. There were more male (n=148, 60%) than female patients and the median age was 64 years (range 18-98 years), with 58% of patients over 60 and 12% under 40 years. The Covid-19 diagnosis was confirmed by a positive SARS-CoV-2 PCR in 204 (89%) patients, and typical thoracic CT results was available in most of the patients (81%). The most frequent symptoms of Covid-19 were fever (76%), cough (63%), dyspnea (60%) and fatigue (50%). One-hundred and fourteen patients (47%) were admitted to the ICU with a median stay of 27 days, and 10 (4%) died. Male gender, non-Caucasian origin, and the presence of comorbidities (obesity, diabetes, cardiopathy, cancer) were associated with a greater Covid-19 severity. Higher age was associated with greater Covid-19 severity but not with ICU admission (Tables 1 and 2).

**Table 1.** Comparison between Covid-19 severity groups.

|  |  | 1. non- | 2. hospitalized |  |  |  |
| --- | --- | --- | --- | --- | --- | --- |
|  | all | hospitalized (a) | (b) | 3. ICU (c) | 4. deaths (d) | p‡ |
|  | N=245 | N=24 (9.8%) | N=96 (39.2%) | N=114 (46.5%) | N=10 (4.1%) |  |
| Age | 63.8<br>[50.2, 72.7] | 42.2<br>[32.3, 55.9] b, c,<br>d | 72.1<br>[62.2, 85.9] a, c | 59.4<br>[50.2, 66.9] a, b,<br>d | 74.1<br>[65.5, 77.0] a, c | <0.001* |
| ageINF40 | 29 (12%) | 11 (48%) b, c, d | 6 (6%) a | 12 (11%) a | 0 (0%) a | <0.001* |
| ageSUP60 | 140 (58%) | 5 (22%) b, d | 72 (76%) a, c | 54 (47%) b | 8 (80%) a | <0.001* |
| Gender (F) | 97 (40%) | 11 (46%) | 48 (50%) c | 34 (30%) b | 4 (40%) | 0.020* |
| Ethnicity |  |  |  |  |  | 0.017* |
| *north African* | 42 (17%) | 3 (12%) | 16 (17%) | 23 (20%) | 0 (0%) |  |
| *sub-Saharan African* | 32 (13%) | 1 (4%) | 12 (12%) | 18 (16%) | 1 (10%) |  |
| *Caribbean* | 5 (2%) | 0 (0%) | 2 (2%) | 3 (3%) | 0 (0%) |  |
| *Asian* | 14 (6%) | 1 (4%) | 2 (2%) | 8 (7%) | 3 (30%) |  |
| *other* | 3 (1%) | 0 (0%) | 1 (1%) | 1 (1%) | 1 (10%) |  |
| *Caucasian* | 92 (38%) | 13 (54%) | 46 (48%) | 29 (25%) | 3 (30%) |  |
| *not available* | 57 (23%) | 6 (25%) | 17 (18%) | 32 (28%) | 2 (20%) |  |
| Caucasian |  |  |  |  |  | 0.008* |
| *no* | 96 (39%) | 5 (21%) c | 33 (34%) c | 53 (46%) a, b | 5 (50%) |  |
| *yes* | 92 (38%) | 13 (54%) | 46 (48%) | 29 (25%) | 3 (30%) |  |
| *not available* | 57 (23%) | 6 (25%) | 17 (18%) | 32 (28%) | 2 (20%) |  |
| BMI | 26.7<br>[22.7, 30.2] | 24.6<br>[21.3, 26.3] c | 25.7<br>[21.9, 29.4] | 27.9<br>[23.3, 31.0] a, d | 22.1<br>[19.9, 25.4] c | 0.002* |
| Comorbidities | 190 (78%) | 9 (39%) b, c | 73 (76%) a | 99 (88%) a | 8 (80%) | <0.001* |
| Cardiac disease | 131 (54%) | 2 (9%) b, c, d | 55 (57%) a | 68 (60%) a | 6 (60%) a | <0.001* |
| Respiratory disease | 26 (11%) | 2 (9%) | 16 (17%) | 7 (6%) | 1 (10%) | 0.089 |
| Diabetes | 70 (29%) | 2 (9%) | 25 (26%) | 39 (35%) | 4 (40%) | 0.045* |
| Active smoking | 39 (16%) | 3 (13%) | 12 (12%) | 22 (19%) | 2 (20%) | 0.528 |
| Obesity | 52 (21%) | 2 (9%) | 12 (12%) c | 37 (33%) b | 1 (10%) | <0.001* |
| Immunodeficiency | 15 (6%) | 0 (0%) | 5 (5%) | 9 (8%) | 1 (10%) | 0.401 |
| Cancer | 11 (5%) | 0 (0%) | 10 (10%) c | 1 (1%) b | 0 (0%) | 0.008* |
| Other comorbidity | 22 (9%) | 1 (4%) | 9 (9%) | 9 (8%) | 2 (20%) | 0.481 |
| Covid-19 Diagnosis by PCR | 204 (89%) | 14 (74%) | 81 (88%) | 98 (92%) | 10 (100%) | 0.121 |
| Covid-19 Diagnosis by CT | 164 (81%) | 8 (67%) | 62 (77%) | 85 (86%) | 8 (89%) | 0.180 |
| Other diagnosis criteria | 42 (17%) | 12 (50%) b, c, d | 18 (19%) a | 12 (11%) a | 0 (0%) a | 0.001* |
| Covid-19 symptoms |  |  |  |  |  |  |
| Fever | 183 (76%) | 18 (75%) | 69 (73%) | 90 (81%) | 6 (60%) | 0.301 |
| Cough | 149 (63%) | 12 (50%) | 52 (55%) | 79 (73%) | 6 (67%) | 0.028* |
| Chest pain | 26 (12%) | 5 (21%) | 7 (8%) | 13 (13%) | 1 (14%) | 0.230 |
| Myalgia | 45 (20%) | 11 (46%) b, c | 15 (17%) a | 17 (17%) a | 2 (29%) | 0.014* |
| Dyspnea | 141 (60%) | 6 (25%) c, d | 42 (45%) c, d | 83 (77%) a, b | 10 (100%) a, b | <0.001* |
| Nausea / Vomiting | 34 (15%) | 5 (21%) | 13 (14%) | 15 (14%) | 1 (12%) | 0.865 |
| Diarrhea | 52 (23%) | 6 (25%) | 17 (19%) | 27 (26%) | 2 (25%) | 0.659 |
| Fatigue | 115 (50%) | 17 (71%) | 52 (55%) | 42 (40%) | 4 (44%) | 0.021* |
| Anosmia | 39 (18%) | 11 (48%) b, c | 10 (11%) a | 17 (17%) a | 1 (17%) | <0.001* |
| Dysgueusia | 28 (13%) | 9 (39%) b, c | 5 (6%) a | 13 (13%) a | 1 (14%) | <0.001* |

Notes. Data are given as median [first, third quartiles] for continuous variables and as count (percentages) for categorical variables.
‡ The Kruskal-Wallis test was used to compare groups for continuous variables and Fisher's Exact test for qualitative variables. Pairwise Wilcoxon-Mann-Whitney tests for continuous variables and pairwise Fisher's Exact tests for qualitative variables were performed using Benjamini-Hochberg method to correct for multiple testing. Letters in brackets indicate which groups significantly differ: (a) group differs from 1.non-hospitalized ; (b) group differs from 2. hospitalized; (c) group differs from 3. ICU; (d) group differs from 4. deaths.
Abbreviations: BMI = body mass index; CT = computed tomography; ECMO = extracorporeal membrane oxygenation; F= female; ICU = intensive care unit; N= number ; NA= Not Applicable.

**Table 2.** Results from adjacent category logit model on Covid-19 severity.

|  | P[Y=2. hospitalized] /<br>P[Y=1.non-hospitalized] |  | P[Y=3. ICU] /<br>P[Y=2. hospitalized] |  | P[Y=4. death] /<br>P[Y=3. ICU] |  |
| --- | --- | --- | --- | --- | --- | --- |
|  | OR [CI95%] | p | OR [CI95%] | p | OR [CI95%] | p |
| Age | 1.12 [1.07-1.16] | <0.001* | 0.95 [0.93-0.97] | <0.001* | 1.06 [1.01-1.12] | 0.018* |
| Gender (M) | 1 [0.32-3.13] | 0.998 | 2 [1.05-3.81] | 0.034* | 0.81 [0.19-3.44] | 0.772 |
| At least one comorbidity (yes) | 3.5 [1.05-11.62] | 0.041* | 1.96 [0.86-4.50] | 0.110 | 0.63 [0.11-3.50] | 0.598 |
| Obesity (yes) | 1.41 [0.23-8.53] | 0.707 | 2.37 [1.05-5.35] | 0.037* | 0.36 [0.04-3.28] | 0.367 |

Notes. For categorical effects, category in brackets is not the reference
Abbreviations: M: male; OR: Odds ratios ; CI: Confidences Intervals; ICU : intensive care unit

### Neurological and psychiatric symptoms and signs

Neuropsychiatric symptoms were very diverse (Figure 1A). The most frequently reported neuropsychiatric symptoms were motor deficit (41%), cognitive disturbances (35%), impaired consciousness (26%), psychiatric disturbance (24%), headache (20%) and behavioral disturbance (18%).

**Figure 1.**
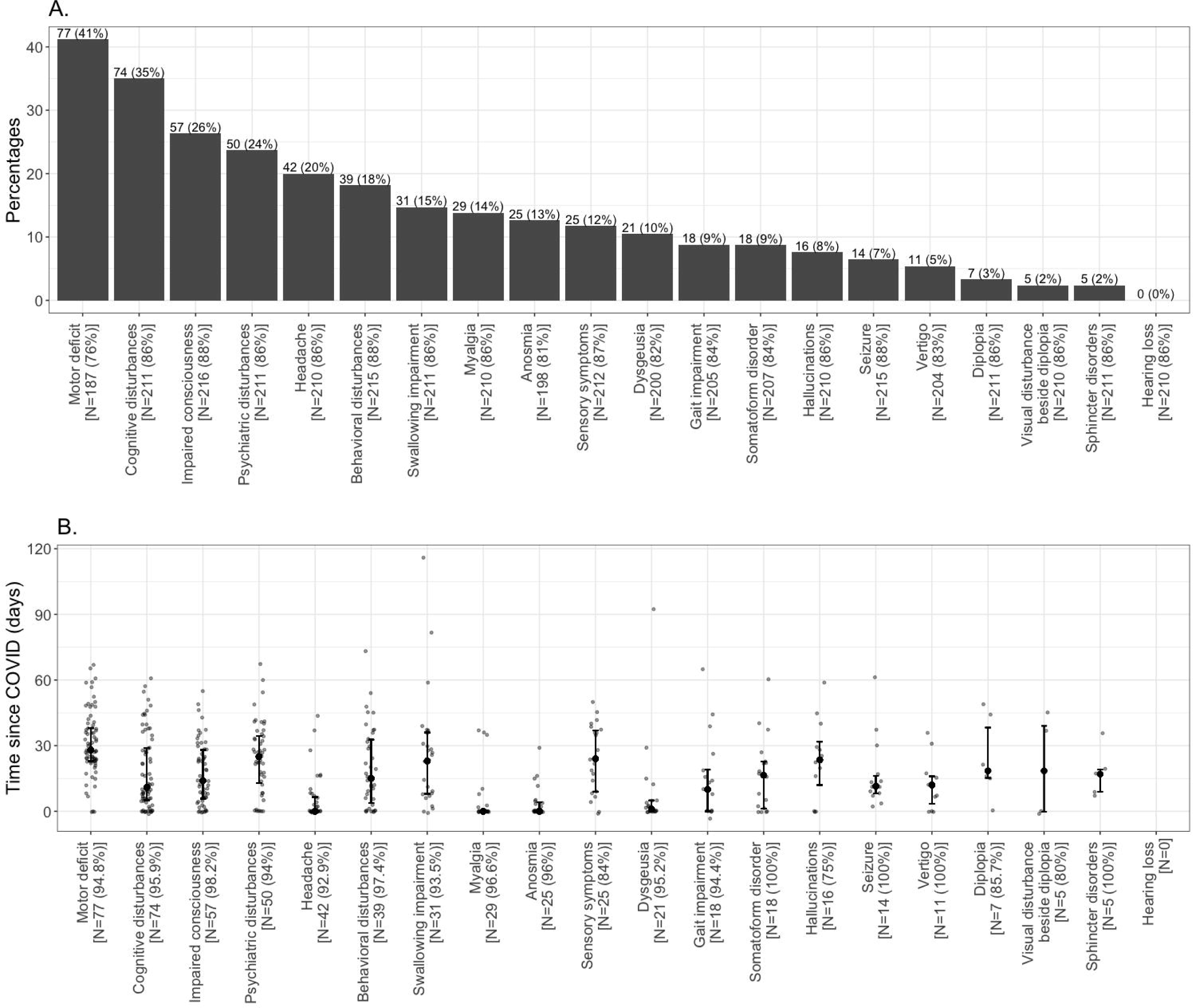
Neuropsychiatric symptoms and their delays since Covid-19 onset. A. Neuropsychiatric symptoms repartition. For each symptom, the number and percentage of non-missing patients is given below. B. Delay between symptom and COVID onset for each symptom. Median, first and third quartiles are represented. The number of subjects with the symptom as well as the percentage of available delays among them are given below.

Psychiatric symptoms were detailed for 29 (12%) of patients. The most frequent psychiatric symptoms were depressed mood (n=13, 48%), agitation (n=11, 38%), loss of appetite (n=9, 32%), social anxiety (n=8, 28%), insomnia and persecution (n=6, 21%, each), anger (n=3, 11%), fear of death (n=4, 14%), and suicidal thoughts (n=3, 11%).

The delay between Covid-19 symptoms onset and the appearance of neuropsychiatric symptoms was highly variable and ranged from 3 to 116 days (Figure 1B). Some symptoms appeared soon after Covid-19 onset: myalgia (median 0 days), headache (0 days), anosmia (0 days), dysgeusia (1 day), gait impairment (10 days). Conversely, some symptoms appeared with a delayed onset, such as sensory symptoms (24 days), psychiatric disturbances (25 days), or motor deficits (28 days).

Clinical examination features are presented in Figure 2. All types of neurological signs were observed, the most frequent being motor deficit (41%), cognitive disturbance (38%) including frontal syndrome (14%), temporo-spatial disorientation (33%), memory (26%) or language (18%) disorders. Cranial nerve examination was abnormal in 21% of the patients (cranial nerve II: 2 patients, III, IV or VI: 15 patients, V: 4 patients, VII: 16 patients, VIII: 2 patients, IX or X: 12, XI: 4 patients, XII: 5 patients), either seen in the context of brainstem global dysfunction or isolated cranial nerve palsies. Movement disorders, mostly myoclonus and myoclonic tremor, were seen in 14% of the patients and cerebellar syndrome in 2%. Pyramidal syndrome was present in only 12% of the patients and areflexia in 10%.

**Figure 2.**
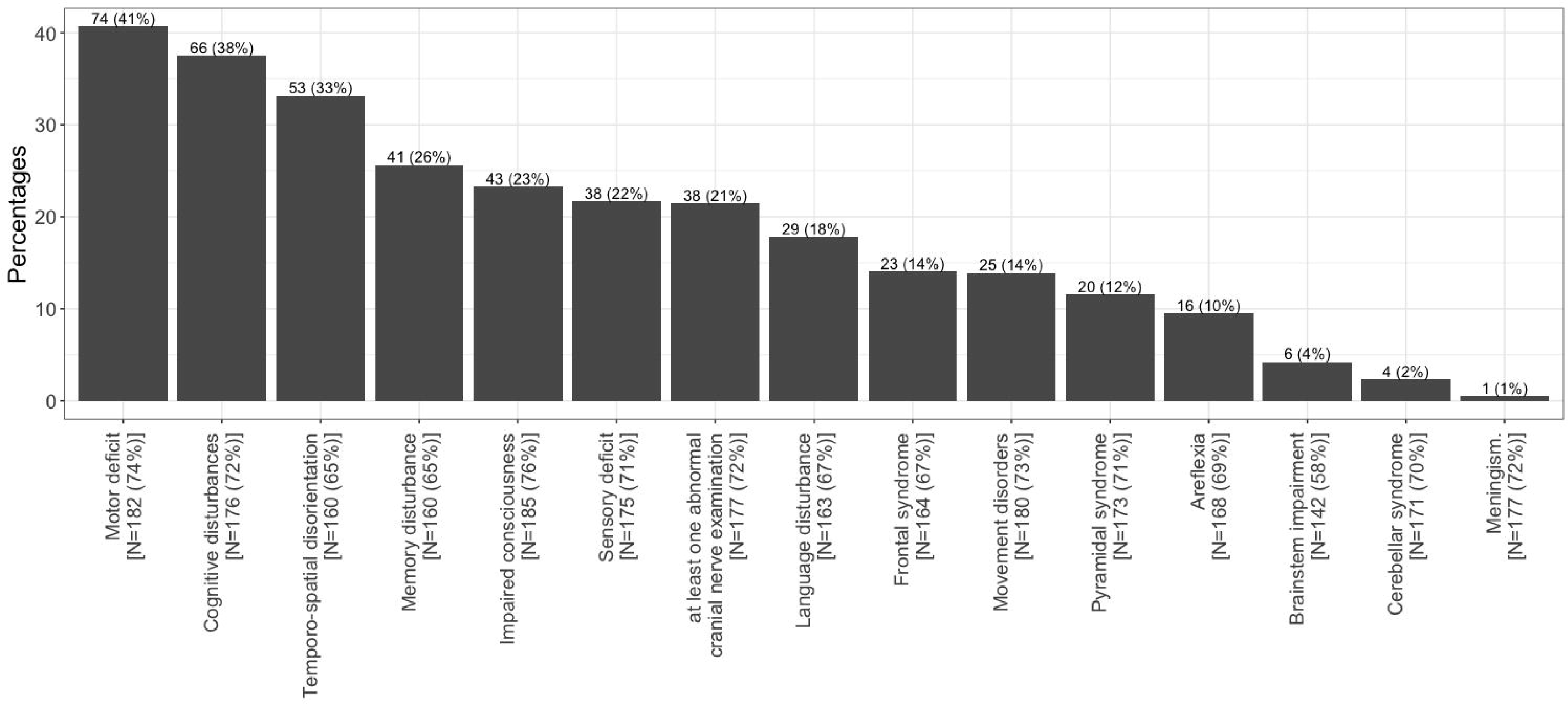
Clinical examination features repartition. For each feature, the number and percentage of non-missing patients is given below.

### Paraclinical explorations

CSF was collected in 53 (22%) patients. Only six patients (11%) showed evidence of CSF hypercellularity (leucocytes >5/mm^3^), ranging from 6 to 205 leucocytes /mm^3^. Protein count was above 0.40g/l in 17 patients (0.42 – 2.9g/l). SARS-CoV-2 PCR was negative in the CSF for all patients in which this analysis was performed (38 patients).

Brain MRI was performed in 119 patients and was abnormal in 81 patients (68%, 9 patients with missing data). In most cases, MRI findings were non-specific and the main features have been described elsewhere ^23^.

Eight patients underwent brain FDG-PET/CT imaging, which was abnormal in six patients. FDG-PET/CT features in some of these patients have been reported elsewhere ^24^. EEG was performed in 82 patients and was abnormal in 54 patients (77%, 12 with missing data). Detailed EEG data is part of another paper (Lambrecq et al., under review). Pathological EEG findings included focal abnormalities, metabolic-toxic encephalopathy, periodic discharge and epileptic activity.

ENMG was carried out in 25 patients and was abnormal in 20 patients with evidence of peripheral nervous system impairment (87%, 2 patients with missing results).

### Syndromes and causes of neuropsychiatric complications

The most frequent syndromes were encephalopathy (42%), critical illness polyneuropathy and myopathy (26%), isolated psychiatric disturbance (18%), and cerebrovascular disorders (16%) (Figure 3A). Other syndromes were much rarer: persistent headache (7%), seizures (6%), isolated movement disorders (4%), cognitive disturbance without encephalopathy (3%), encephalitis (3%). Guillain-Barré syndrome was observed in five patients, and isolated cranial nerve impairment in five patients. Two patients had posterior reversible encephalopathy syndrome (PRES). One patient had myelitis confirmed on spine MRI. Three patients displayed a cerebellar syndrome. Three patients had signs of brainstem impairment. Four patients complained of subjective sensory signs without ENMG abnormalities. Two patients showed somatoform disorders.

**Figure 3.**
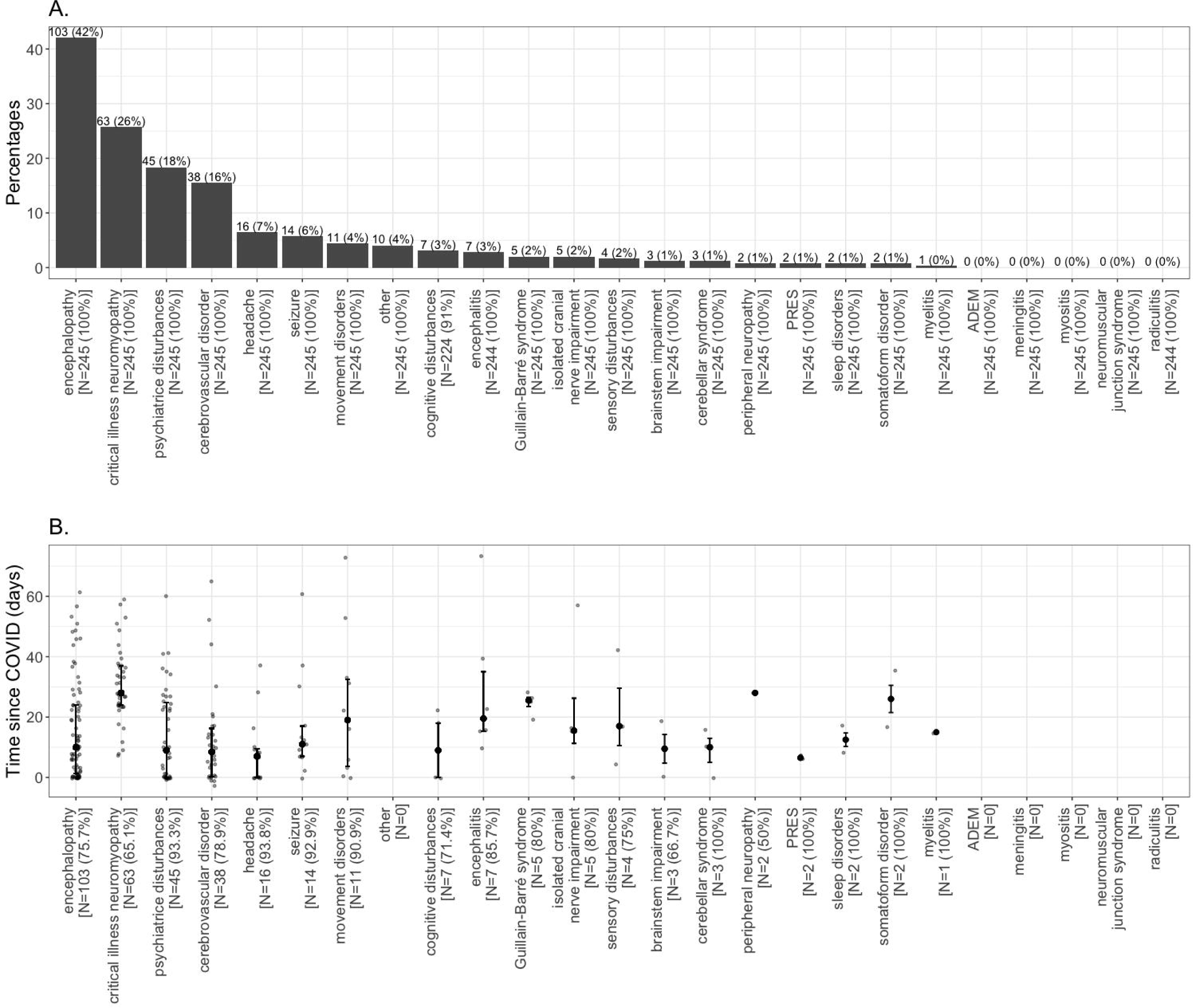
Neuropsychiatric syndromes and their delays since Covid-19 onset. A. Neuropsychiatric syndromes repartition. For each syndrome, the number and percentage of non-missing patients is given below. B. Delay between syndrome and COVID onset for each syndrome. Median, first and third quartiles are represented. The number of subjects with the syndrome as well as the percentage of available delays among them are given below.

The delay between Covid-19 onset and each neuropsychiatric syndrome onset is presented in Figure 3B. A wide range of delay was seen across and within each diagnosis, with several trends. Cerebrovascular disorders, cognitive disturbance, headache and psychiatric disturbance usually occurred within the first ten days following Covid-19 onset. Conversely, myelitis, encephalitis and cranial nerve palsies occurred around 15 days, Guillain-Barré and somatoform disorders around 25 days, and critical-illness neuromyopathy after 28 days from Covid-19 onset.

### Encephalopathy

Among patients with encephalopathy, 56% were hospitalized in the ICU. Twenty percent of patients with encephalopathy had a pre-existing neurological disorder, mostly neurodegenerative disorders with pre-existing dementia. Seventy percent of the patients with encephalopathy had at least one cardiological, respiratory or metabolic comorbidity. The most common clinical presentations of encephalopathy were confusion, ICU-delirium, and delayed awakening after stopping sedative drugs.

Among the ten deceased patients, seven (70%) showed encephalopathy. With respect to risk ratios, encephalopathy mainly concerned patients over 60 (Table 3). In the correlation matrix, encephalopathy was associated with higher age, death, cardiac and diabetic comorbidities (Figure 4A).

**Table 3.** Risk ratios for each potential risk factor of the 3 most frequent syndromes.

| Potential risk factors | Critical illness |  |  |
| --- | --- | --- | --- |
|  | neuromyopathy | Cerebrovascular disorder | Encephalopathy |
|  | N=63 | N=38 | N=106 |
| Gender (female) (n=97) | 0.61 (n=18) | 0.70 (n=12) | **0.78 (n=36)** |
| No comorbidities (n=58) | 0.47 (n=8) | **0.60 (n=6)** | **0.66 (n=18)** |
| Comorbidity |  |  |  |
| *cardiac disorder (n=131)* | 0.96 (n=33) | **2.44 (n=28)** | 1.18 (n=61) |
| *obesity (n=52)* | 2.13 (n=23) | 0.84 (n=7) | 0.81 (n=19) |
| *diabetes (n=70)* | 1.44 (n=23) | 1.30 (n=13) | 1.29 (n=36) |
| Age |  |  |  |
| *age < 40 (n=31)* | 1.00 (n=8) | 0.81 (n=4) | 0.20 (n=3) |
| *40 < age < 60 (n=74)* | 1.73 (n=27) | 1.51 (n=15) | 0.96 (n=31) |
| *60 < age < 80 (n=105)* | 1.07 (n=28) | 0.87 (n=15) | 1.33 (n=31) |
| *80 < age (n=35)* | 0.00 (n=0) | 0.71 (n=4) | 1.31 (n=19) |
| ICU (N=114) | **22.98 (n=60)** | 0.41 (n=10) | 1.29 (n=56) |

Notes: In bold, the significant risk factors.
Abbreviations: ICU = intensive care unit.

**Figure 4.**
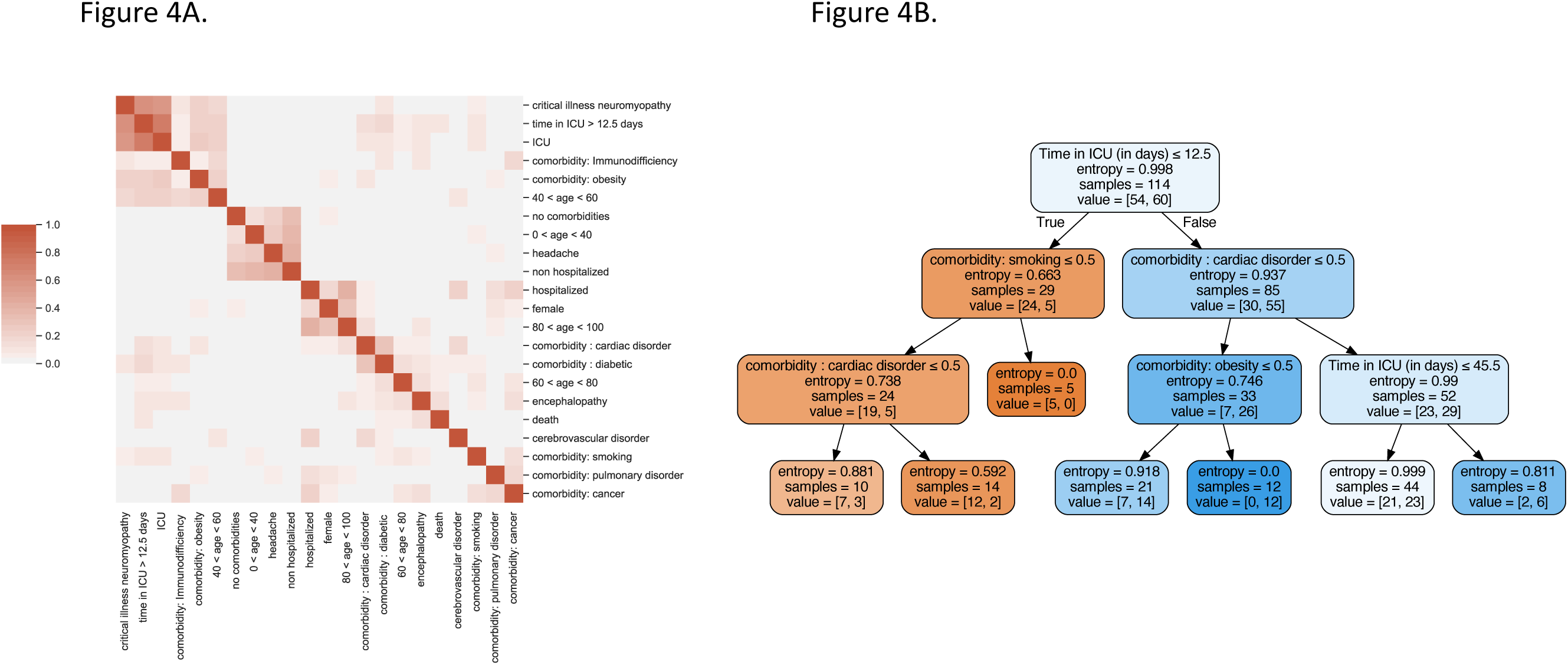
Statistical associations between neuropsychiatric syndromes and risk factors. A. Correlation matrix between neuropsychiatric syndromes, comorbidities and other related variables. Shades of red indicates a positive correlation. The variables were clustered by minimizing the single linkage distance. B. Decision tree trained to predict whether a patient who went to ICU had an ICU neuropathy or not. The depth of the tree was imposed to 4 and trained through entropy minimization. 114 patients went to ICU and 60 had an ICU neuropathy.

### Critical illness polyneuropathy and myopathy

Critical illness neuropathy or myopathy was diagnosed in the recovery phase after sedative drug reduction in the ICU. ENMG findings were available for 22% of these patients. Several patterns of neuromuscular injury were observed: i) axonal sensorimotor polyneuropathy, ii) myopathy, iii) troncular nerve compressions (median, ulnar, peroneal, lateral femoral cutaneous nerves), iv) brachial plexopathy. Brachial plexopathy was only seen in patients after remaining prone for extended periods. Thirty-seven percent of the patients with critical illness polyneuropathy had pre-existing diabetes.

The decision tree showed that a stay exceeding 12.5 days was the strongest feature predictive of a neuropathy after entering the ICU (Figure 4B).

### Psychiatric disorders

Among the 28 patients with available details for their psychiatric disturbances, 71% had a pre-existing psychiatric disorder (depression (70%), anxiety disorders (20%), psychosis (20%), bipolar disorder (10%), substance abuse disorder (5%)).

The most commonly observed psychiatric disorders in the context of Covid-19 infection were anxiety disorders (25%), depression (18%), psychosis (20%), adjustment disorders (7%) and acute stress (3%).

### Cerebrovascular disorders

Thirty-eight patients (16%, median [Q1,Q3] age 62.3 [52.1,70.1]; 26 males, 68%) suffered strokes with the following proportions: 32 (84%) ischemic strokes (regional or multiple small infarcts), three (8%) parenchymal hematomas, one subarachnoid hemorrhage and one cerebral venous thrombosis. Patients with cerebrovascular disorders had the following cardiovascular risk factors: hypertension (22, 58%), diabetes mellitus (13, 34%), dyslipidemia (9, 24%) and obesity with body mass index ≥30 (7, 18%). The correlation matrix indeed showed the association between cerebrovascular disorders and comorbidities (smoking, pulmonary disorder, cancer) (Figure 4A). Seven patients had a history of previous stroke. Unusual symptoms at presentation (which were not explained by infarct location or metabolic disturbances) were frequently found such as delirium (13/32, 40%) or apathy (5/38, 13%).

### Seizures

Among the 14 patients with seizures, none had a previous history of epilepsy. One patient was under treatment for Parkinson’s disease with dementia and one patient for glioblastoma. Three patients had a focal seizure without generalization, one patient a focal to bilateral generalized seizure, seven patients a generalized seizure, one patient a status epilepticus.

### Cranial nerve palsies

Five patients presented with cranial nerve palsies. One patient had VI^th^ nerve palsy with normal MRI and CSF examination. As she was a heavy smoker, a thrombosis triggered by the Covid-19 infection was suspected. One patient presented with optic neuritis, in the setting of a possible inflammatory disorder of the CNS (inflammatory lesions on brain MRI, oligoclonal bands in the CSF). One patient presented with III^rd^ nerve partial palsy after an ICU stay. One patient presented with unilateral hypoglossal nerve palsy, and one with combined homolateral X, XI and XII nerves palsies, while in the ICU, which were attributed to mechanical complications of positioning, intubation, or jugular catheterizations.

### Headaches

Isolated major headache was the primary diagnosis in 16 patients (7%). Headaches often had migraine characteristics. None of the patients had pre-existing migraines. Among patients with headache, eight underwent brain MRI, which was normal in seven patients and showed a non-specific lesion in one. Two had lumbar puncture (normal in both). None of the patients had evidence of meningitis or intracranial hypertension. In the correlation matrix, headache is associated with younger patients who were not hospitalized and showed no associated comorbidities (Figure 4A).

### Encephalitis

Only seven patients fulfilled criteria for encephalitis. Among those seven patients, one patient had positive PCR for varicella-zoster-virus (VZV) in the CSF, suggesting concomitant VZV encephalitis. One patient had an encephalopathy with myoclonus and inflammatory brain lesions, one patient had cognitive disturbances with myoclonus and CSF pleiocytosis, one patient had brainstem impairment, movement disorders and dysautonomia with white matter lesions on MRI, two patients showed alterations of consciousness with white matter lesions on MRI, one patient had confusion with MRI features of limbic encephalitis. Several of these patients have been already reported ^25^.

### Guillain-Barré

Five patients were diagnosed with Guillain-Barré syndrome, three of them being hospitalized in the ICU. One patient had a pre-existing demyelinating Charcot-Marie-Tooth disease. In four patients, Guillain-Barré syndrome presented with severe motor deficit of all four limbs. One of the patients had associated bilateral facial nerve palsy. CSF showed elevated protein in two patients (1.1g/l and 1.65 g/l). Four patients underwent ENMG, which showed typical characteristics of acute inflammatory demyelinating polyneuropathy in all of them.

## Discussion

We have described a wide range of neuropsychiatric symptoms and syndromes occurring in Covid-19 patients seen in a single multidisciplinary center over a six month period. The major categories of neuropsychiatric manifestations in our cohort were: encephalopathy, critical illness polyneuropathy and myopathy, psychiatric disturbance, and cerebrovascular complications.

The prevalence of neuropsychiatric symptoms among patients seen in our Covid-19 center was 12%. The prevalence of neurological signs reported in previous studies has been very variable, depending on inclusion criteria and evaluation methods, varying between 4.2% ^26^ and 57.4% ^27^. The relatively lower prevalence in our study may be due to the strict inclusion criteria and the exclusion of patients with isolated symptoms of anosmia or dysgeusia. Our cohort of patients was comparable to previous cohorts in terms of mean age, clinical severity, gender and comorbidities. Compared to those with non-severe disease, the patients in ICU were more likely to have comorbidities, including hypertension, diabetes, cancer, cardiac or kidney disease.

Our cohort highlights the high frequency of neuropsychiatric complications related to critical illness and intensive care. Almost half of the patients in our cohort were hospitalized in the ICU (47%). Our findings emphasize that encephalopathy is a major issue in patients with Covid-19. The presence of encephalopathy was strongly associated with Covid-19 infection severity and the presence of comorbidities, which is in keeping with complications of hypoxia and critical illness. The high prevalence of encephalopathy in patients with Covid-19 patients has already been reported, though its mechanisms are not fully understood. Covid-19 encephalopathy seems to be more common in patients with more severe Covid-19-related respiratory disease, associated comorbidities, evidence of multi-organ system dysfunction, including hypoxemia, renal and hepatic impairment, and elevated markers of systemic inflammation ^28–30^. The association of encephalopathy with greater age and comorbidities has already been reported ^31^. A recent large-scale cohort also emphasized the frequency of non-specific complications, including toxic/metabolic encephalopathy and hypoxic/ischemic brain injury ^32^. It is noteworthy that encephalopathy was associated with the risk of death in our cohort.

Critical care polyneuropathy and myopathy were particularly frequent in our patients, affecting 26% of them. Besides classical ICU polyneuropathy, many patients presented with mechanic plexopathy and nerve compressions, probably secondary to prolonged sedation and lying prone, which was usual in the Covid-19 patients with acute respiratory distress syndrome. The long duration of critical care, the requirement for high doses of anesthetics and curare, and the frequent association with diabetes could partly explain the particularly high frequency of ICU neuropathy compared to what is usually seen with other viral respiratory infections. Two patients presented with mechanical cranial nerve injuries (XII in one patient; X, XI and XII in one patient) in the setting of critical care, a complication known as Tapia syndrome ^33^.

Acute cerebrovascular disorders consisted mostly in ischemic strokes. The risk of cerebrovascular disorders was highly associated with the presence of comorbidities. The presence of unusual stroke symptoms at presentation, such as encephalopathy without focal deficit, is noteworthy and was also reported by other authors^34^. Covid-19 might facilitate ischemic strokes via at least three non-exclusive pathogenic mechanisms: atherosclerotic plaque vulnerability, a hypercoagulable state, and cerebral microvasculature injury (endothelitis). Indeed, Covid-19 is commonly complicated by sepsis-induced coagulopathy, induced by a systemic inflammatory response involving endothelial dysfunction and microthrombosis often associated with multi organ failure ^35^.

These observations of frequent unspecific neurological features contrast with the rarity of syndromes potentially linked to viral neuroinvasion. The question of the neuroinvasiveness of the SARS-CoV-2 is an ongoing debate. Its detection in the CSF has only been reported in a few case studies ^6,7^. Only seven patients in our cohort fulfilled the diagnostic criteria for encephalitis ^21^. A recent cohort with 606 patients with neurological signs of Covid-19 reported no encephalitis ^32^, suggesting this is a very rare complication. None of the patients with encephalitis had evidence of SARS-CoV-2 detection in the CSF. The CSF pleiocytosis without SARS-CoV-2 detection and brain inflammatory lesions seen in our patients and other reported cases of Covid-19 might result from immune-mediated inflammatory mechanisms rather than direct viral invasion. The role of cytokine activation has been widely reported for Covid-19 ^36,37^. Headaches are also proposed to be based on an inflammatory cytokinic mechanism ^38^. Psychiatric manifestations could also be related to inflammatory mechanisms ^39^. Neuropathological studies have, to date, failed to demonstrate specific alterations and the majority of neuropathological observations favor the role of critical illness complications ^40,41^. A recent neuropathological study found evidence of inflammatory brain lesions, with no evidence of brain damage directly caused by the SARS-CoV-2 ^42^.

Other immune-mediated complications, such as Guillain-Barré syndrome and myelitis occurred anecdotally in our cohort. It should be stressed that Guillain-Barré syndrome and myelitis had a delayed onset, which is in keeping with a post-infectious inflammatory mechanism. Cranial nerve impairments could often be explained by alternative mechanisms (probable pre-existing inflammatory disorder in one patient, thrombotic mechanism in another and mechanical compressions in a further two).

Although our sample of patients who developed psychiatric symptoms during Covid-19 is small, our findings are in accordance with previously reported data ^43^. Indeed, our patients mainly suffered from anxiety disorder or depression, and one developed *de novo* psychosis, obsessive compulsive disorder or bipolar disorder. Long-term follow-up data will be valuable to have an estimation of the risk of developing post-traumatic stress disorder, especially for patients with adjustment disorders and acute stress syndrome.

Our study has several limitations. Although the study is retrospective, it was implemented early at the onset of the Covid-19 pandemic and data could be collected prospectively by the investigators as they examined new patients. We may have an over representation of complications due to critical illness and critical care as there was a large number of Covid-19-dedicated ICU beds in our institution. We cannot exclude reporting bias and lack of exhaustivity, but the involvement of all physicians of the Department implicated in every step of patient care (ICU, acute hospitalization departments, rehabilitation departments, imaging departments, neurophysiology) limits this bias, especially for the most severe forms of neuropsychiatric manifestations. In stroke patients, one limitation of the study is that we could not determine for all patients the etiology of ischemic strokes and in case of a negative outcome, whether Covid-19 was the trigger.

## Conclusion

We report the broad landscape of neuropsychiatric complications in a large cohort of Covid-19 patients. The majority of these complications could be imputed to critical illness, intensive care and systemic inflammation, which contrasts with the paucity of more direct SARS-CoV2-related complications or post-infectious disorders. Further studies are needed to better disentangle the different mechanisms underlying these various symptoms, and to explore potential long-term complications.

## Acknowledgement

The Cohort COVID-19 Neurosciences (CoCo Neurosciences) study supported by the APHP and funded by the generous support of the FIA (Fédération Internationale pour l’Automobile) Foundation and donors of the Paris Brain Institute – ICM. The authors thank the CoCo-Neurosciences study group for their participation in the data collection. The research leading to these results received funding from the program “*Investissements d’avenir*” ANR-10-IAIHU-06.

The Coco-Neurosciences study group: **Steering Committee** (Pitié-Salpêtrière Hospital, Paris): Cecile Delorme, Jean-Christophe Corvol, Jean-Yves Delattre, Stephanie Carvalho, Sandrine Sagnes. **Scientific Committee** (Pitié-Salpêtrière Hospital, Paris): Bruno Dubois, Vincent Navarro, Celine Louapre, Tanya Stojkovic, Ahmed Idbaih, Charlotte Rosso, David Grabli, Ana Zenovia Gales, Bruno Millet, Benjamin Rohaut, Eleonore Bayen, Sophie Dupont, Gaelle Bruneteau, Stephane Lehericy, Danielle Seilhean, Alexandra Durr, Foudil Lamari, Marion Houot, Vanessa Batista Brochard. **Principal investigators**: <u>Pitié-Salpêtrière Hospital (Paris)</u>: Sophie Dupont, Catherine Lubetzki, Danielle Seilhean, Pascale Pradat-Diehl, Charlotte Rosso, Khe Hoang-Xuan, Bertrand Fontaine, Lionel Naccache, Philippe Fossati, Isabelle Arnulf, Alexandra Durr, Alexandre Carpentier, Stephane Lehericy, Yves Edel; <u>Rothschild Hospital (Paris)</u>: Gilberte Robain, Philippe Thoumie; <u>Avicenne Hospital (Bobigny)</u>: Bertrand Degos; <u>Sainte-Anne Hospital (Paris)</u>: Tarek Sharshar; <u>Saint-Antoine Hospital (Paris)</u>: Sonia Alamowitch, Emmanuelle Apartis-Bourdieu, Charles-Siegried Peretti; <u>Saint-Louis Hospital (Paris)</u>: Renata Ursu; <u>Tenon Hospital (Paris)</u>: Nathalie Dzierzynski; <u>Charles Foix Hospital (Ivry)</u>: Kiyoka Kinugawa Bourron, Joel Belmin, Bruno Oquendo, Eric Pautas, Marc Verny. **Co-investigators**: <u>Pitié-Salpêtrière Hospital (Paris)</u>: Cecile Delorme, Jean-Christophe Corvol, Jean-Yves Delattre, Yves Samson, Sara Leder, Anne Leger, Sandrine Deltour, Flore Baronnet, Ana Zenovia Gales,Stephanie Bombois, Mehdi Touat, Ahmed Idbaih, Marc Sanson, Caroline Dehais, Caroline Houillier, Florence Laigle-Donadey, Dimitri Psimaras, Agusti Alenton, Nadia Younan, Nicolas Villain, David Grabli, Maria del Mar Amador, Gaelle Bruneteau, Celine Louapre, Louise-Laure Mariani, Nicolas Mezouar, Graziella Mangone, Aurelie Meneret, Andreas Hartmann, Clement Tarrano, David Bendetowicz, Pierre-François Pradat, Michel Baulac, Sara Sambin, Phintip Pichit, Florence Chochon, Adele Hesters, Bastien HerlinAn Hung Nguyen, Valerie Procher, Alexandre Demoule, Elise Morawiec, Julien Mayaux, Morgan Faure, Claire Ewenczyk, Giulia Coarelli, Anna Heinzmann, Tanya Stojkovic, Marion Masingue, Guillaume Bassez, Vincent Navarro, Isabelle An, Yulia Worbe, Virginie Lambrecq, Rabab Debs, Esteban Munoz Musat, Timothee Lenglet, Virginie Lambrecq, Aurelie Hanin, Lydia Chougar, Nathalia Shor, Nadya Pyatigorskaya, Damien Galanaud, Delphine Leclercq, Sophie Demeret, Benjamin Rohaut, Albert Cao, Clemence Marois, Nicolas Weiss, Salimata Gassama, Loic Le Guennec, Vincent Degos, Alice Jacquens, Thomas Similowski, Capucine Morelot-Panzini, Jean-Yves Rotge, Bertrand Saudreau, Bruno Millet, Victor Pitron, Nassim Sarni, Nathalie Girault, Redwan Maatoug, Ana Zenovia Gales, Smaranda Leu, Eleonore Bayen, Lionel Thivard, Karima Mokhtari, Isabelle Plu; <u>Sainte-Anne Hospital (Paris)</u>: Bruno Gonçalves; <u>Saint-Antoine Hospital (Paris)</u>: Laure Bottin, Marion Yger; <u>Rothschild Hospital (Paris)</u>: Gaelle Ouvrard, Rebecca Haddad; <u>Charles Foix Hospital (Ivry)</u>: Flora Ketz, Carmelo Lafuente, Christel Oasi. **Other Contributors**: **Associated centers** (Lariboisière Hospital, Paris): Bruno Megabarne, Dominique Herve; **Clinical Research Associates** (ICM, Pitié-Salpêtrière Hospital, Paris): Haysam Salman, Armelle Rametti-Lacroux, Alize Chalançon, Anais Herve, Hugo Royer, Florence Beauzor, Valentine Maheo, Christelle Laganot, Camille Minelli, Aurelie Fekete, Abel Grine, Marie Biet, Rania Hilab, Aurore Besnard, Meriem Bouguerra, Gwen Goudard, Saida Houairi, Saba Al-Youssef, Christine Pires, Anissa Oukhedouma, Katarzyna Siuda-Krzywicka, Tal Seidel Malkinson; (Saint-Louis Hospital, Paris): Hanane Agguini; **Data Manager** (ICM, Paris): Safia Said; **Statistician** (ICM, Paris): Marion Houot.

## Funding

The study received funding from the FIA (*Federation Internationale de l’Automobile*.)

## Financial disclosures

Cécile Delorme received a research grant from the FIA, travel funding from Merz Pharma, Boston Scientific and Medtronic.

Vincent Navarro served as board member for UCB pharma, LivaNova, GW pharma et EISAI. Louise-Laure Mariani has received research support grants from INSERM, JNLF, The L’Oreal Foundation; speech honoraria from CSL, Sanofi-Genzyme, Lundbeck, Teva; consultant for Alzprotect and received travel funding from the Movement Disorders Society, ANAINF, Merck, Merz, Medtronic, Teva and AbbVie, outside the submitted work.

J.C. Corvol served on the scientific advisory boards for Biogen, UCB, Prevail Therapeutic, Idorsia, Ever Pharma, Denali, and has received grants from the Michael J Fox Foundation and Sanofi outside of this work.

The other authors declare no potential financial conflict.

## Appendix 1. Author contributions

| Name | Location | Contribution |
| --- | --- | --- |
| Cécile Delorme | APHP, Paris | Design, data acquisition, data analysis and interpretation, drafted and revised the manuscript |
| Marion Houot | APHP, Paris | Data analysis and interpretation, revised the manuscript for intellectual content |
| Charlotte Rosso | APHP, Paris | Data acquisition, revised the manuscript for intellectual content |
| Stéphanie Carvalho | APHP, Paris | Data acquisition, revised the manuscript for intellectual content |
| Thomas Nedelec | APHP, Paris | Data analysis and interpretation, revised the manuscript for intellectual content |
| Redwan Maatoug | APHP, Paris | Data acquisition, revised the manuscript for intellectual content |
| Victor Pitron | APHP, Paris | Data acquisition, revised the manuscript for intellectual content |
| Salimata Gassama | APHP, Paris | Data acquisition, revised the manuscript for intellectual content |
| Sara Sambin | APHP, Paris | Data acquisition, revised the manuscript for intellectual content |
| Stéphanie Bombois | APHP, Paris | Data acquisition, revised the manuscript for intellectual content |
| Bastien Herlin | APHP, Paris | Data acquisition, revised the manuscript for intellectual content |
| Gaëlle Ouvrard | APHP, Paris | Data acquisition, revised the manuscript for intellectual content |
| Gaëlle Bruneteau | APHP, Paris | Data acquisition, revised the manuscript for intellectual content |
| Adèle Hesters | APHP, Paris | Data acquisition, revised the manuscript for intellectual content |
| Ana Zenovia Gales | APHP, Paris | Data acquisition, revised the manuscript for intellectual |
|  |  | content |
| Bruno Millet | APHP, Paris | Data acquisition, revised the manuscript for intellectual content |
| Foudil Lamari | APHP, Paris | Data acquisition, revised the manuscript for intellectual content |
| Stéphane Lehericy | APHP, Paris | Data acquisition, revised the manuscript for intellectual content |
| Vincent Navarro | APHP, Paris | Data acquisition, revised the manuscript for intellectual content |
| Benjamin Rohaut | APHP, Paris | Data acquisition, revised the manuscript for intellectual content |
| Sophie Demeret | APHP, Paris | Data acquisition, revised the manuscript for intellectual content |
| Thierry Maisonobe | APHP, Paris | Data acquisition, revised the manuscript for intellectual content |
| Marion Yger | APHP, Paris | Data acquisition, revised the manuscript for intellectual content |
| Bertrand Degos | APHP, Paris | Data acquisition, revised the manuscript for intellectual content |
| Louise-Laure Mariani | APHP, Paris | Data acquisition, revised the manuscript for intellectual content |
| Christophe Bouche | APHP, Paris | Data acquisition, revised the manuscript for intellectual content |
| Nathalie Dzierzynski | APHP, Paris | Data acquisition, revised the manuscript for intellectual content |
| Bruno Oquendo | APHP, Paris | Data acquisition, revised the manuscript for intellectual content |
| Flora Ketz | APHP, Paris | Data acquisition, revised the manuscript for intellectual content |
| An-Hung Nguyen | APHP, Paris | Data acquisition, revised the manuscript for intellectual content |
| Aurélié Kas | APHP, Paris | Data acquisition, revised the manuscript for intellectual content |
| Jean-Yves Delattre | APHP, Paris | Design, data acquisition, |
|  |  | revised the manuscript for intellectual content |
| Jean-Christophe Corvol | APHP, Paris | Design, data acquisition, revised the manuscript for intellectual content |

## Notes

### Competing Interest Statement

Cecile Delorme received a research grant from the FIA, travel funding from Merz Pharma, Boston Scientific and Medtronic. 
Vincent Navarro served as board member for UCB pharma, LivaNova, GW pharma et EISAI. 
Louise-Laure Mariani has received research support grants from INSERM, JNLF, The L Oreal Foundation; speech honoraria from CSL, Sanofi-Genzyme, Lundbeck, Teva; consultant for Alzprotect and received travel funding from the Movement Disorders Society, ANAINF, Merck, Merz, Medtronic, Teva and AbbVie, outside the submitted work.
J.C. Corvol served on the scientific advisory boards for Biogen, UCB, Prevail Therapeutic, Idorsia, Ever Pharma, Denali, and has received grants from the Michael J Fox Foundation and Sanofi outside of this work.
The other authors declare no potential financial conflict. 

### Clinical Trial

NCT04362930

### Author Declarations

The study was conducted in accordance with good clinical practice, the French regulation for retrospective studies on clinical data, and was compliant with the European General Data Protection Regulation (GDPR) and the French Commission Nationale de l Informatique et des Libertes (CNIL) rules. All patients (or their relatives in cases of impaired consciousness) received written information about the research, and consented to the use of their data. The study received the approval of the Sorbonne University Ethics Committee (N 2020 CER-202028). The study is registered on the clinicaltrials.gov website (NCT04362930).

## References

1. Helms J, Kremer S, Merdji H, et al. Neurologic Features in Severe SARS-CoV-2 Infection. N Engl J Med. Epub 2020 Apr 15.:NEJMc2008597.

2. Hemasian H, Ansari B. First case of Covid-19 presented with cerebral venous thrombosis: A rare and dreaded case. Revue Neurologique. Epub 2020 May.:S0035378720305579.

3. Muhammad S, Petridis A, Cornelius JF, Hänggi D. Letter to editor: Severe brain haemorrhage and concomitant COVID-19 Infection: A neurovascular complication of COVID-19. Brain Behav Immun. 2020;87:150–151.

4. Oxley TJ, Mocco J, Majidi S, et al. Large-Vessel Stroke as a Presenting Feature of Covid-19 in the Young. N Engl J Med. Epub 2020 Apr 28.:e60.

5. Chougar L, Mathon B, Weiss N, Degos V, Shor N. Atypical Deep Cerebral Vein Thrombosis with Hemorrhagic Venous Infarction in a Patient Positive for COVID-19. AJNR Am J Neuroradiol. 2020;41:1377–1379.

6. Moriguchi T, Harii N, Goto J, et al. A first Case of Meningitis/Encephalitis associated with SARS-Coronavirus-2. International Journal of Infectious Diseases. Epub 2020 Apr.:S1201971220301958.

7. Virhammar J, Kumlien E, Fällmar D, et al. Acute necrotizing encephalopathy with SARS-CoV-2 RNA confirmed in cerebrospinal fluid. Neurology. Epub 2020 Jun 25.:10.1212/WNL.0000000000010250.

8. Abdi S, Ghorbani A, Fatehi F. The association of SARS-CoV-2 infection and acute disseminated encephalomyelitis without prominent clinical pulmonary symptoms. Journal of the Neurological Sciences. 2020;416:117001.

9. Le Guennec L, Devianne J, Jalin L, et al. Orbitofrontal involvement in a neuroCOVID-19 patient. Epilepsia. Epub 2020 Jun 26.:epi.16612.

10. Vollono C, Rollo E, Romozzi M, et al. Focal status epilepticus as unique clinical feature of COVID-19: A case report. Seizure. 2020;78:109–112.

11. Zachariadis A, Tulbu A, Strambo D, Dumoulin A, Di Virgilio G. Transverse myelitis related to COVID-19 infection. J Neurol [online serial]. Epub 2020 Jun 29. Accessed at: http://link.springer.com/10.1007/s00415-020-09997-9. Accessed July 11, 2020.

12. Zhao H, Shen D, Zhou H, Liu J, Chen S. Guillain-Barré syndrome associated with SARS-CoV-2 infection: causality or coincidence? The Lancet Neurology. Epub 2020 Apr.:S1474442220301095.

13. Dinkin M, Gao V, Kahan J, et al. COVID-19 presenting with ophthalmoparesis from cranial nerve palsy. Neurology. Epub 2020 May 1.:10.1212/WNL.0000000000009700.

14. Gutiérrez-Ortiz C, Méndez A, Rodrigo-Rey S, et al. Miller Fisher Syndrome and polyneuritis cranialis in COVID-19. Neurology. Epub 2020 Apr 17.:10.1212/WNL.0000000000009619.

15. Laurendon T, Radulesco T, Mugnier J, et al. Bilateral transient olfactory bulbs edema during COVID-19-related anosmia. Neurology. Epub 2020 May 22.:10.1212/WNL.0000000000009850.

16. Politi LS, Salsano E, Grimaldi M. Magnetic Resonance Imaging Alteration of the Brain in a Patient With Coronavirus Disease 2019 (COVID-19) and Anosmia. JAMA Neurol [online serial]. Epub 2020 May 29. Accessed at: https://jamanetwork.com/journals/jamaneurology/fullarticle/2766765. Accessed June 1, 2020.

17. Xydakis MS, Dehgani-Mobaraki P, Holbrook EH, et al. Smell and taste dysfunction in patients with COVID-19. The Lancet Infectious Diseases. Epub 2020 Apr.:S1473309920302930.

18. Rogers JP, Chesney E, Oliver D, et al. Psychiatric and neuropsychiatric presentations associated with severe coronavirus infections: a systematic review and meta-analysis with comparison to the COVID-19 pandemic. The Lancet Psychiatry. Epub 2020 May.:S2215036620302030.

19. Xiong W, Kwan P, Zhou D, Del Felice A, Duncan JS, Sander JW. Acute and late neurological complications of COVID19: the quest for evidence. Brain [online serial]. Epub 2020 Sep 30. Accessed at: https://academic.oup.com/brain/advance-article/doi/10.1093/brain/awaa294/5913080. Accessed October 15, 2020.

20. WHO. Coronavirus Disease (COVID-19) Situation Reports [online]. 2020. Accessed at: https://www.who.int/emergencies/diseases/novel-coronavirus-2019/situation-reports. Accessed June 28, 2020.

21. Ellul MA, Benjamin L, Singh B, et al. Neurological associations of COVID-19. The Lancet Neurology. Epub 2020 Jul.:S1474442220302210.

22. Pedregosa F, Varoquaux G, Gramfort A, et al. Scikit-learn: Machine Learning in Python. MACHINE LEARNING IN PYTHON. :6.

23. Chougar L, Shor N, Weiss N, et al. Retrospective Observational Study of Brain Magnetic Resonance Imaging Findings in Patients with Acute SARS-CoV-2 Infection and Neurological Manifestations. Radiology. Epub 2020 Jul 17.:202422.

24. Delorme C, Paccoud O, Kas A, et al. Covid-19-related encephalopathy: a case series with brain FDG-PET/CT findings. European Journal of Neurology [online serial]. n/a. Accessed at: https://onlinelibrary.wiley.com/doi/abs/10.1111/ene.14478. Accessed August 16, 2020.

25. Cao A, Rohaut B, Guennec LL, et al. Severe COVID-19-related encephalitis can respond to immunotherapy. Brain [online serial]. Accessed at: https://academic.oup.com/brain/advance-article/doi/10.1093/brain/awaa337/5926056. Accessed October 19, 2020.

26. Xiong W, Mu J, Guo J, et al. New onset neurologic events in people with COVID-19 infection in three regions in China. Neurology. Epub 2020 Jun 17.:10.1212/WNL.0000000000010034.

27. Romero-Sánchez CM, Díaz-Maroto I, Fernández-Díaz E, et al. Neurologic manifestations in hospitalized patients with COVID-19: The ALBACOVID registry. Neurology. Epub 2020 Jun 1.:10.1212/WNL.0000000000009937.

28. Mao L, Wang M, Chen S, et al. Neurological Manifestations of Hospitalized Patients with COVID-19 in Wuhan, China: a retrospective case series study [online]. Infectious Diseases (except HIV/AIDS); 2020 Feb. Accessed at: http://medrxiv.org/lookup/doi/10.1101/2020.02.22.20026500. Accessed April 1, 2020.

29. Pinna P, Grewal P, Hall JP, et al. Neurological manifestations and COVID-19: Experiences from a tertiary care center at the Frontline. Journal of the Neurological Sciences. 2020;415:116969.

30. Radnis C, Qiu S, Jhaveri M, Da Silva I, Szewka A, Koffman L. Radiographic and clinical neurologic manifestations of COVID-19 related hypoxemia. Journal of the Neurological Sciences. 2020;418:117119.

31. Liotta EM, Batra A, Clark JR, et al. Frequent neurologic manifestations and encephalopathy-associated morbidity in Covid-19 patients. Annals of Clinical and Translational Neurology [online serial]. Epub 2020 Oct 5. Accessed at: https://onlinelibrary.wiley.com/doi/10.1002/acn3.51210. Accessed October 15, 2020.

32. Frontera JA, Sabadia S, Lalchan R, et al. A Prospective Study of Neurologic Disorders in Hospitalized COVID-19 Patients in New York City. Neurology. Epub 2020 Oct 5.:10.1212/WNL.0000000000010979.

33. Decavel P, Petit C, Tatu L. Tapia syndrome at the time of the COVID-19 pandemic: Lower cranial neuropathy following prolonged intubation. Neurology. 2020;95:312–313.

34. Katz JM, Libman RB, Wang JJ, et al. Cerebrovascular Complications of COVID-19. Stroke [online serial]. Epub 2020 Aug 6. Accessed at: https://www.ahajournals.org/doi/10.1161/STROKEAHA.120.031265. Accessed August 18, 2020.

35. Coolen T, Lolli V, Sadeghi N, et al. Early postmortem brain MRI findings in COVID-19 non-survivors. Neurology. Epub 2020 Jun 16.:10.1212/WNL.0000000000010116.

36. Muccioli L, Pensato U, Cani I, Guarino M, Cortelli P, Bisulli F. COVID-19– Associated Encephalopathy and Cytokine-Mediated Neuroinflammation. Ann Neurol. 2020;88:860–861.

37. Ye Q, Wang B, Mao J. The pathogenesis and treatment of the ‘Cytokine Storm’ in COVID-19. Journal of Infection. Epub 2020 Apr.:S0163445320301651.

38. Membrilla JA, Lorenzo í, Sastre M, Díaz de Terán J. Headache as a Cardinal Symptom of Coronavirus Disease 2019: A Cross-Sectional Study. Headache: The Journal of Head and Face Pain. Epub 2020 Sep 28.:head.13967.

39. Guo Q, Zheng Y, Shi J, et al. Immediate psychological distress in quarantined patients with COVID-19 and its association with peripheral inflammation: A mixed-method study. Brain, Behavior, and Immunity. 2020;88:17–27.

40. Deigendesch N, Sironi L, Kutza M, et al. Correlates of critical illness-related encephalopathy predominate postmortem COVID-19 neuropathology. Acta Neuropathol [online serial]. Epub 2020 Aug 26. Accessed at: http://link.springer.com/10.1007/s00401-020-02213-y. Accessed September 12, 2020.

41. Pezzini A, Padovani A. Lifting the mask on neurological manifestations of COVID-19. Nat Rev Neurol [online serial]. Epub 2020 Aug 24. Accessed at: http://www.nature.com/articles/s41582-020-0398-3. Accessed August 29, 2020.

42. Matschke J, Lütgehetmann M, Hagel C, et al. Neuropathology of patients with COVID-19 in Germany: a post-mortem case series. The Lancet Neurology [online serial]. Epub 2020 Oct. Accessed at: https://linkinghub.elsevier.com/retrieve/pii/S1474442220303082. Accessed October 12, 2020.

43. Holmes EA, O’Connor RC, Perry VH, et al. Multidisciplinary research priorities for the COVID-19 pandemic: a call for action for mental health science. The Lancet Psychiatry. Epub 2020 Apr.:S2215036620301681.

